# Adaptive immune responses to SARS-CoV-2 in recovered severe COVID-19 patients

**DOI:** 10.1101/2021.01.05.20249027

**Authors:** Beatriz Olea, Eliseo Albert, Ignacio Torres, Paula Amat, María José Remigia, Roberto Gozalbo-Rovira, Jesús Rodríguez-Díaz, Javier Buesa, María Luisa Blasco, Josep Redón, Jaime Signes-Costa, David Navarro

**Author notes:** **Correspondence:** David Navarro, Microbiology Service, Hospital Clínico Universitario, Instituto de Investigación INCLIVA, Valencia, and Department of Microbiology, University of Valencia, Valencia, Spain. Av. Blasco Ibáñez 17, 46010 Valencia, Spain.

## Abstract

**Objectives:** There is an imperative need to determine the durability of adaptive immunity to SARS-CoV-2. We enumerated SARS-CoV-2-reactive CD4^+^ and CD8^+^ T cells targeting S1 and M proteins and measured RBD-specific serum IgG over a period of 2-6 months after symptoms onset in a cohort of subjects who had recovered from severe clinical forms of COVID-19.

**Methods:** We recruited 58 patients (38 males and 20 females; median age, 62.5 years), who had been hospitalized with bilateral pneumonia, 60% with one or more comorbidities. IgG antibodies binding to SARS-CoV-2 RBD were measured by ELISA. SARS-CoV-2-reactive CD69^+^-expressing-IFNγ-producing-CD4^+^ and CD8^+^ T cells were enumerated in heparinized whole blood by flow cytometry for ICS.

**Results:** Detectable SARS-CoV-2-S1/M-reactive CD69^+^-IFN-γ CD4^+^ and CD8^+^ T cells were displayed in 17 (29.3%) and 6 (10.3%) subjects respectively, at a median of 84 days after onset of symptoms (range, 58-191 days). Concurrent comorbidities increased the risk (OR, 3.15; 95% CI, 1.03-9.61; *P*=0.04) of undetectable T-cell responses in models adjusted for age, sex and hospitalization ward. Twenty-one out of the 35 patients (60%) had detectable RBD-specific serum IgGs at a median of 118 days (range, 60 to 145 days) after symptoms onset. SARS-CoV-2 RBD-specific IgG serum levels were found to drop significantly over time.

**Conclusion:** A relatively limited number of subjects who developed severe forms of COVID-19 had detectable SARS-CoV-2-S1/M IFNγ CD4^+^ and CD8^+^ T cells at midterm after clinical diagnosis. Our data also indicated that serum levels of RBD-specific IgGs decline over time, becoming undetectable in some patients.

## INTRODUCTION

Experimental evidence supports a major role of neutralizing antibodies (NtAb) and skewed Th1 functional immune responses in preventing and controlling SARS-CoV-2 infection [1-4]. Epitopes eliciting NtAb have been mapped within all SARS-CoV-2 structural proteins; among these, NtAbs targeting the receptor binding domain (RBD) of the viral spike protein (S) appear to display maximum specificity and potency [5-7]. Broad specificity to structural and non-structural proteins has been reported across SARS-CoV-2-reactive CD4^+^ and CD8^+^ T cells, of which S, membrane (M) and nucleocapsid (N) proteins are immunodominant in most individuals [8-18]. Both SARS-CoV-2-specific NtAb and T cells are readily detectable in a large proportion of acute or short-term convalescent COVID-19 patients [8-18], although the strength of adaptive immune responses may be modulated by the severity of the disease [1-4]. Data on SARS-CoV infection suggest that memory B and T cells have a potential for long-lasting persistence (over years) [19,20], yet the durability of SARS-CoV-2 adaptive immunity remains to be established. Determining whether SARS-CoV-2 B- and T-cell responses persist over time following natural infection or after vaccination seems of paramount relevance in designing effective public health policies to prevent virus transmission and spread. In the current study we enumerated SARS-CoV-2-reactive CD4^+^ and CD8^+^ T cells targeting S and M proteins, and measured IgG antibodies binding to RBD of S protein, along a timeframe of up to 6 months after symptoms onset in a cohort of recovered COVID-19 patients who had been hospitalized due to severe clinical forms of the disease.

## PATIENTS AND METHODS

### Patients and specimens

A total of 58 non-consecutive patients (38 males and 20 females; median age, 62.5 years; range, 27 to 82 years) were recruited at the COVID-19 follow-up unit of Hospital Clínico Universitario of Valencia, at a median of 85 days (range, 58 to 191 days) after onset of COVID-19 symptoms. SARS-CoV-2 infection was diagnosed by RT-PCR (between February 26 and May 16) [21]. The only patient inclusion criterion was the availability of serum and/or whole blood specimens for B- and T-cell immunity analyses described below. Medical history and laboratory data were retrospectively reviewed. Clinical severity of COVID-19 was graded following World Health Organization criteria [22]. Whole blood was also collected from seven non-SARS-CoV-2-exposed healthy individuals (up to March 2020) who served as controls. The current study was approved by the Research Ethics Committee of Hospital Clínico Universitario INCLIVA (March 2020).

### SARS-CoV-2 RBD IgG immunoassay

IgG antibodies binding to SARS-CoV-2 RBD made in Sf9 cells infected with recombinant baculoviruses (Invitrogen, CA, USA) were measured by an enzyme-linked immunosorbent assay (ELISA) as previously described [23].

### SARS-CoV-2-reactive IFN-γ CD4^+^ and CD8^+^ T cells

SARS-CoV-2-reactive CD69^+^-expressing-IFNγ-producing-CD4^+^ and CD8^+^ T cells were enumerated in heparinized whole blood by flow cytometry for intracellular cytokine staining (ICS) (BD Fastimmune, BD Biosciences, San Jose, CA, USA) as previously described [21]. Two sets of 15-mer overlapping peptides (11 mer overlap) encompassing the SARS-CoV-2 Spike glycoprotein N-terminal 1-643 amino acid sequence (158 peptides) and the entire sequence of SARS-CoV-2 M protein (53 peptides), were used in combination for stimulation (1□μg/ml per peptide) during 6 h, in the presence of CD28 and CD49d costimulatory mAbs and brefeldin A (10µg/ml), the latter after two hour incubation. Peptide mixes were obtained from JPT Peptide Technologies GmbH (Berlin, Germany). The appropriate positive (phytohemagglutinin) and isotype controls were used. The total number of SARS-CoV-2-reactive CD8^+^ T cells was calculated by multiplying the percentages of CD4^+^ or CD8^+^ T cells producing IFNγ on stimulation (after background subtraction) by the absolute CD4^+^ or CD8^+^ T-cell counts. Responses ≥0.1% were considered specific [21].

### Laboratory measurements

Clinical laboratory investigation included serum levels IL-6, ferritin and Dimer-D, which were monitored at least twice weekly during hospital stay.

### Statistical methods

Frequency comparisons for categorical variables were carried out using the Fisher exact test. Differences between medians were compared using the Mann–Whitney U-test. The Spearman’s rank test was used for analysis of correlation between continuous variables. For logistic regression analyses, variables with *P* values <0.1 in univariate models were included in multivariate models. Two-sided exact *P*-values were reported. A *P*-value <0.05 was considered statistically significant. The analyses were performed using SPSS version 20.0 (SPSS, Chicago, IL, USA).

## RESULTS

### Patient clinical features

All 58 patients in this cohort developed severe forms of COVID-19 requiring hospitalization either in the intensive care unit (ICU) (n=21) or in other hospital wards (n=37). All patients presented with bilateral pneumonia and 60% had one or more comorbidities, including diabetes mellitus, asthma, hypertension, dyslipidemia, cancer or chronic lung disease. All ICU patients underwent mechanical ventilation. Median hospitalization of patients was 16 days (range, 6-61 days). Patients in the two groups were matched for age, sex and comorbidities (not shown).

### SARS-CoV-2-reactive IFN-γ CD4^+^ and CD8^+^ T cells in recovered COVID-19 patients

SARS-CoV-2-S1/M-reactive CD69^+^-IFN-γ CD4^+^ and CD8^+^ T cells were enumerated at a median of 84 days after symptoms onset (range, 58-191 days). Representative plots are shown in Supplementary Figure 1. Of the 58 patients, 17 (29.3%) and 6 (10.3%) had detectable SARS-CoV-2 CD4^+^ and CD8^+^ T-cell responses, respectively. Only two patients displayed both SARS-CoV-2-reactive T-cell subsets. SARS-CoV-2 CD4^+^ and CD8^+^ T-cell counts ranged from 0.98 to 43.75 cells/µL, and from 0.48 to 2.98 cells/µL, respectively (median, 4.83 and 1.13 cells/µL, respectively).

Figure 1A shows SARS-CoV-2 T-cell reactivity according to the sampling timeframe after symptoms onset (arbitrarily categorized as 2-3 months vs. > 3 months). Overall, we found no difference between the percentage of patients with or without detectable CD4^+^ (*P*=0.40) or CD8^+^ (*P*=0.12) T cells across comparison groups; nevertheless, none of the patients sampled beyond day 130 after COVID-19 presentation (n=3) exhibited either SARS-CoV-2 CD4^+^ or CD8^+^ T-cell responses. Of note, patients with or without detectable SARS-CoV-2-reactive T cells were monitored within a comparable timeframe (median, 91 days; range, 60 to 118 days vs. median 83 days, range, 58 to 191 days; *P*=0.18).

**Figure 1.**
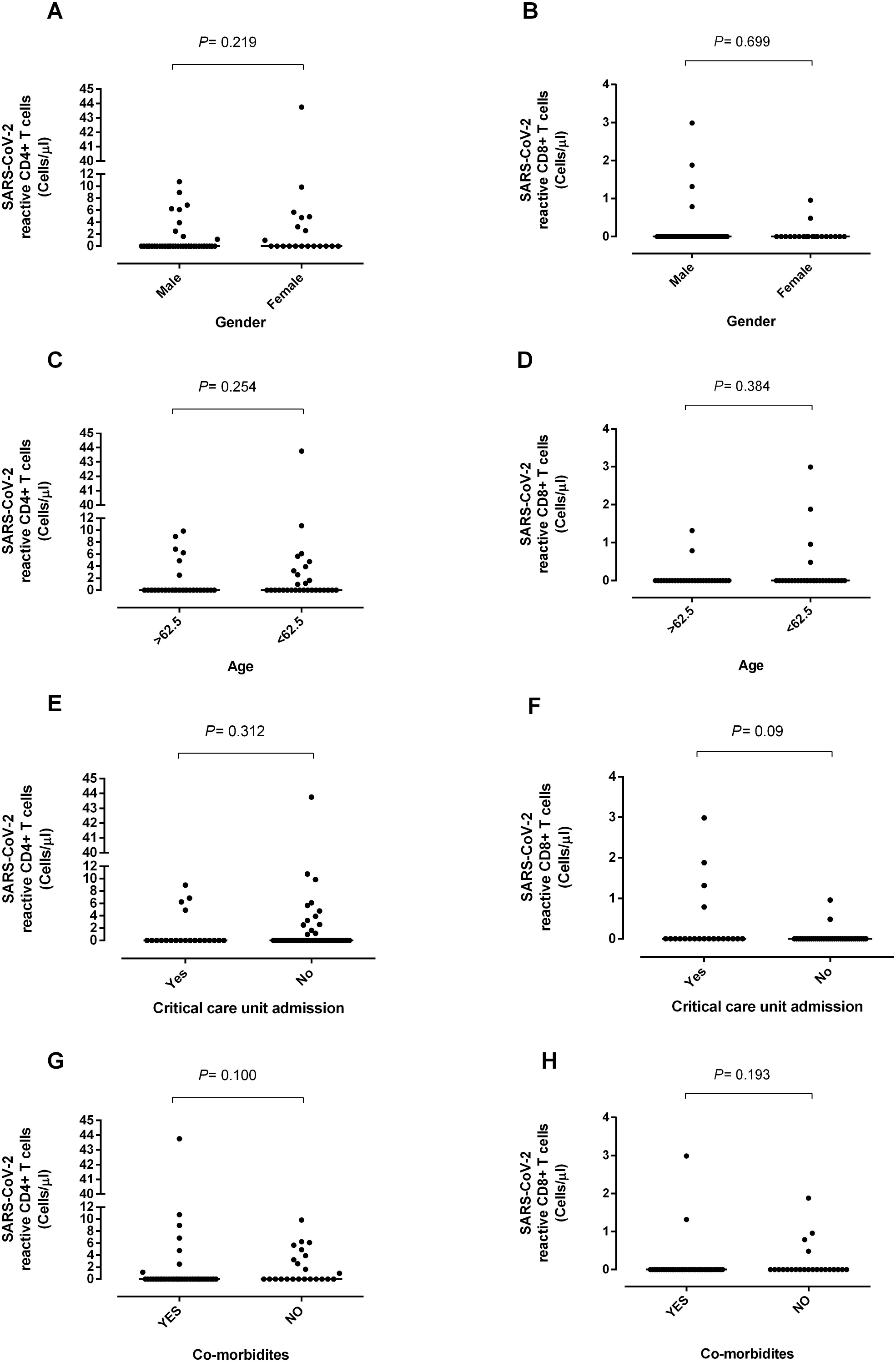
SARS-CoV-2 T- and B-cell responses in individuals recovered from severe COVID-19. Peripheral blood SARS-CoV-2-S1/M-reactive CD69^+^-expressing IFN-γ-producing CD4^+^ and CD8^+^ T cells (A) and SARS-CoV-2-RBD-specific IgG levels (B) according to the time of sampling following symptoms onset. Bars indicate median levels. *P* values are shown.

### SARS-CoV-2 RBD-specific IgGs in recovered COVID-19 patients

Serum specimens for quantitation of SARS-CoV-2 RBD-specific IgGs were available from 35 patients, and collected at a median of 118 days (range, 60 to 145 days) from onset of symptoms. Twenty-one of the 35 patients (60%) had detectable RBD-specific IgGs (median, 1.8 AU/mL; range, 0.99-4.14 AU/mL). RBD-specific IgG reactivity according to time of sampling (2-3 months vs. >3 months) is shown in Figure 1B. A comparable number of patients had detectable responses at both time points (*P*=0.84).

Out of the 21 patients exhibiting RBD-specific IgG reactivity, 10 (47.6%) had measurable SARS-CoV-2 T-cell responses (CD4^+^ T cells in 6 patients and CD8^+^ T cells in the remainder). Of the 14 patients lacking RBD-specific IgGs, 5 had detectable SARS-CoV-2 CD4^+^ T cells and none had SARS-CoV-2 CD8^+^ T cells.

Eighteen of the 35 patients had paired serum samples collected at the time of hospitalization (median, 22 days after symptoms onset; range, 8-34 days) and after recovery (median, 120 days after symptoms onset; range 93-145 days); paired specimens were analyzed in parallel. SARS-CoV-2 RBD-specific IgGs were detectable in 17 patients at the first time point and 14 at the latter. As shown in Figure 2, SARS-CoV-2 RBD-specific IgG serum levels were found to drop significantly over time (from a median of 4.97 AU/ml to a median of 1.51 AU/ml; *P*<0.001).

**Figure 2.**
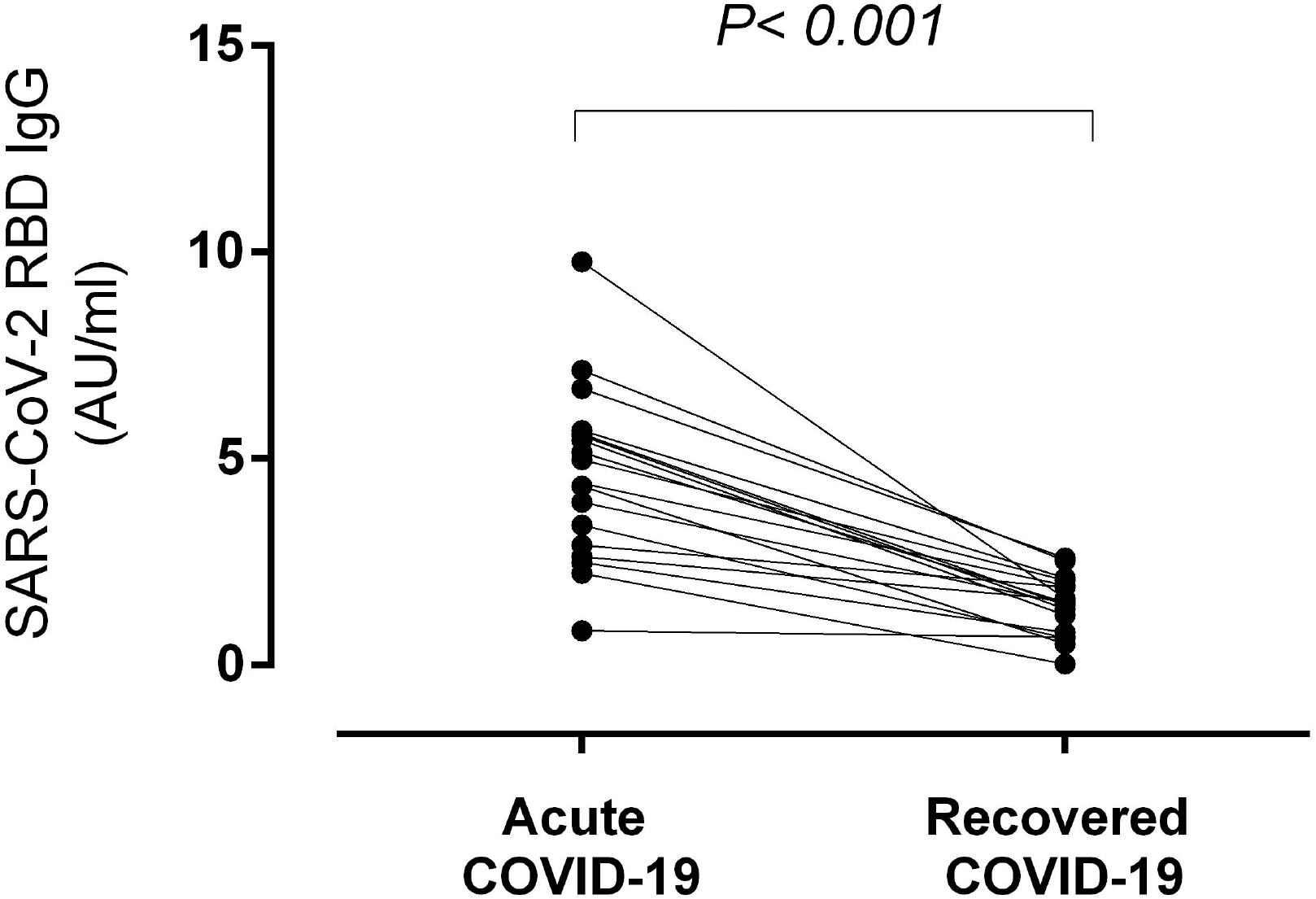
Kinetics of SARS-CoV-2-RBD-specific IgG levels in individuals recovered from severe COVID-19. Serum levels of such an antibody specificity were measured at the time of hospitalization (acute COVID-19) and 2-5 months afterwards (recovered COVID-19). *P* value is shown.

Finally, we found no correlation between SARS-CoV-2 RBD-specific IgG levels and SARS-CoV-2 CD4^+^ and CD8^+^ T-cell counts (*P*=0.12 and *P*=0.14, respectively) in recovered COVID-19 patients.

### Demographic, clinical and biological factors associated with detectable SARS-CoV-2-reactive T cells or RBD-specific IgGs in recovered COVID-19 patients

The detection rate of SARS-CoV-2-reactive T cells (either CD4^+^, CD8^+^ or both) was comparable across patients admitted to ICU or other medical wards (*P*=0.82) (Table 1). Likewise, both median SARS-CoV-2 CD4^+^ and CD8^+^ T-cell counts were similar between groups (Figure 3). Neither age nor sex was found to influence either the likelihood (Table 1) or magnitude (Figure 2) of detectable SARS-CoV-2 T-cell responses. In contrast, patients displaying one or more comorbidities were significantly (*P*=0.04) less likely to exhibit detectable T-cell responses (Table 1), although median T-cell counts were not significantly dissimilar across comparison groups (Figure 3). In fact, comorbidities increased the risk (OR, 3.15; 95% CI, 1.03-9.61; *P*=0.04) of undetectable T-cell responses in logistic regression models adjusted for age, sex and hospitalization ward (ICU vs. others).

**Figure 3.**
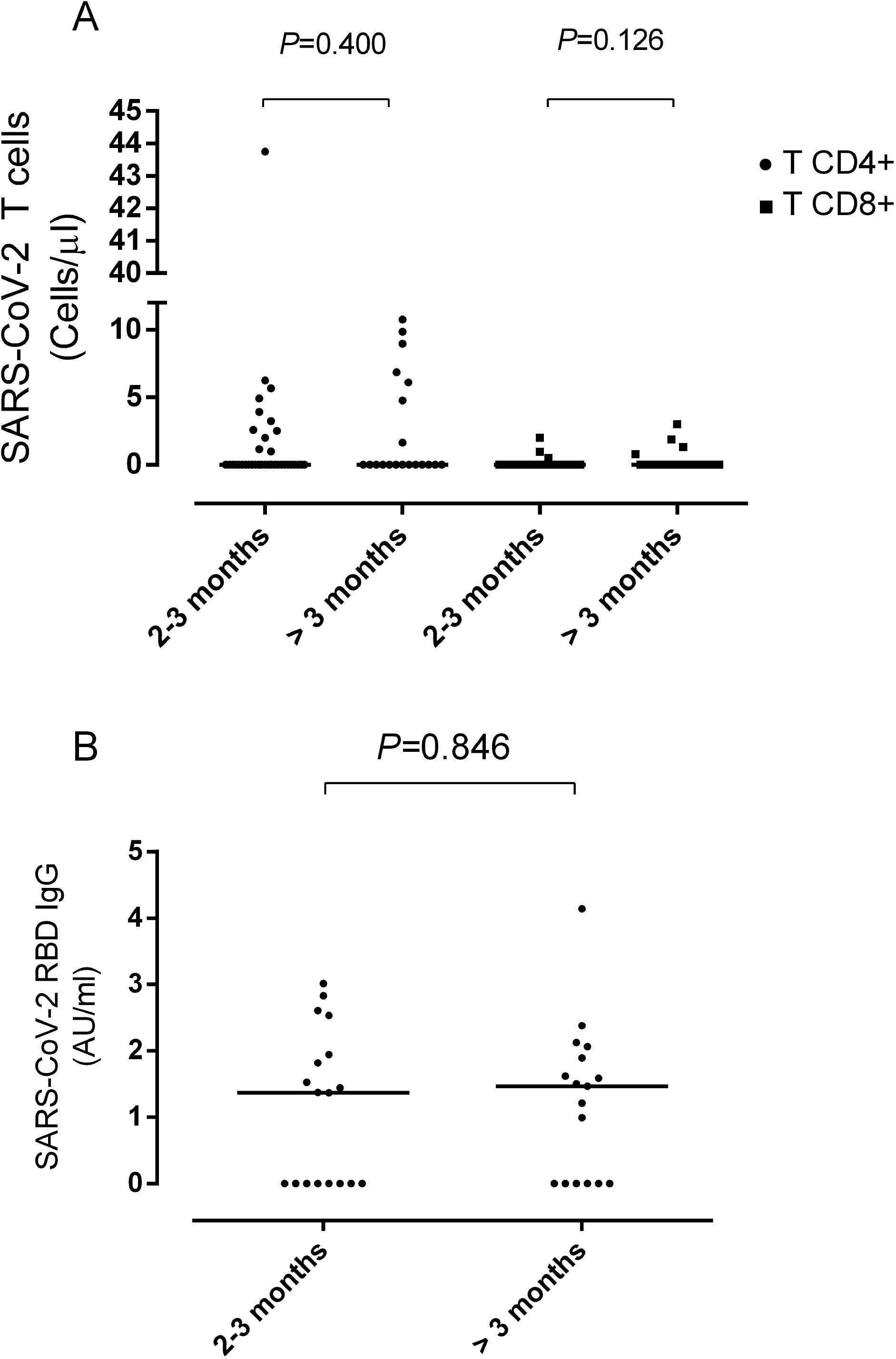
Impact of demographics, hospitalization ward and comorbidities on peripheral blood SARS-CoV-2-S1/M-reactive CD69^+^-expressing IFN-γ-producing CD4^+^ and CD8^+^ T cells in subjects recovered from severe COVID-19. Bars indicate median levels. *P* values are shown.

The likelihood of detecting SARS-CoV-2-RBD-specific IgGs was not influenced by age (*P*=0.67), or presence of comorbidities (*P*=0.65), but was higher in males (*P*=0.004) and in ICU patients (*P*=0.019) (Table 1). Both factors were found to increase the likelihood of detecting an antibody response in multivariate models adjusted for age and comorbidities (OR, 11.71; 95% CI, 1.86-73.7; *P*=0.009 and OR, 6.57; 95% CI, 1.04-41.4; *P*=0.04, respectively). Nevertheless, neither of these parameters had a significant impact on serum SARS-CoV-2 IgG levels (not shown).

The net state of inflammation shortly after viral infections may shape the quality and strength of ongoing adaptive immunity responses [24]. In this context, we next compared serum peak levels of several inflammatory biomarkers, including IL-6, ferritin and Dimer-D, measured within the first 15 days after hospitalization among recovered COVID-19 patients with or without detectable SARS-CoV-2 T-cell responses. As shown in Figure 4, no significant differences were found between comparison groups for any of these parameters. A similar observation was made for SARS-RBD-specific IgGs (not shown). Furthermore, was observed either weak or no correlation between serum peak levels of IL-6, ferritin or Dimer-D and SARS-CoV-2 CD4^+^ and CD8^+^ T-cell counts or SARS-CoV-2 RBD-specific IgG levels (Supplementary Table 1).

**Figure 4.**
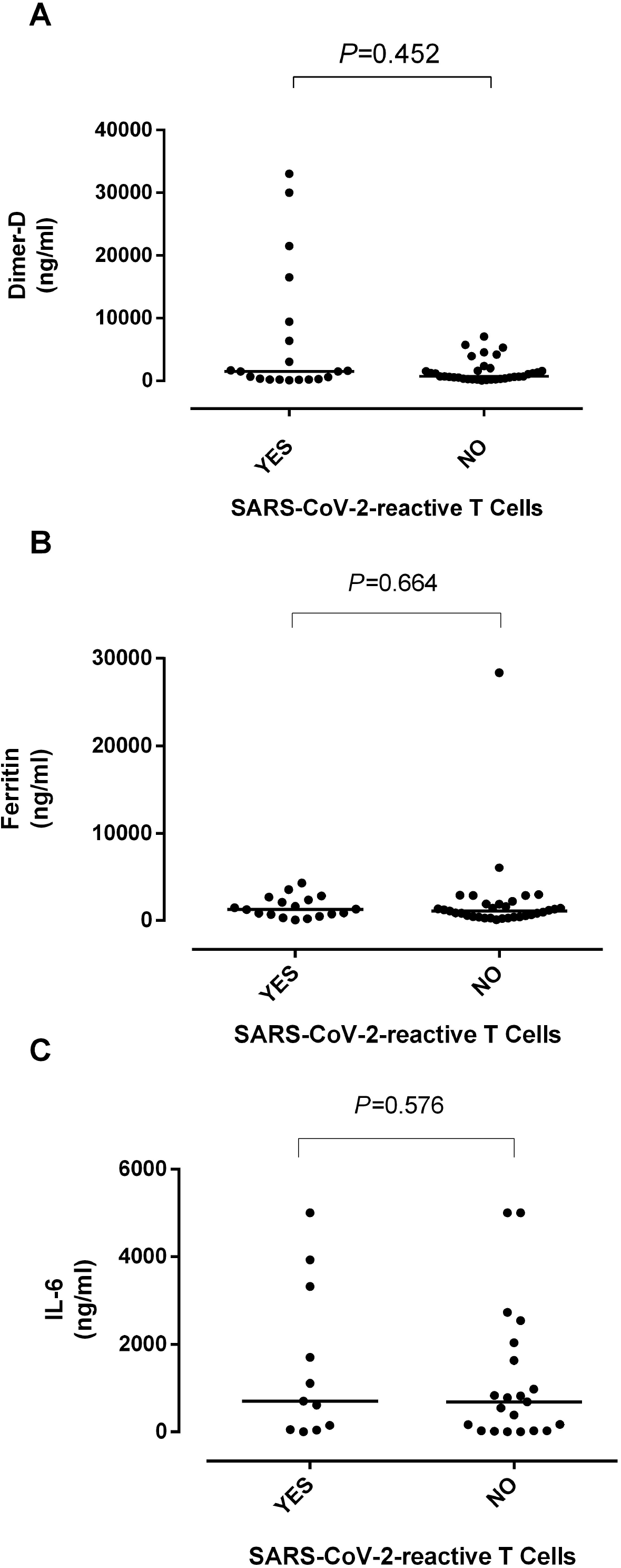
Serum peak levels of inflammatory biomarkers (IL-6, Ferritin and Dimer-D) measured within the first 15 days after hospitalization and SARS-CoV-2-S1/M-reactive CD69^+^-expressing IFN-γ-producing CD4^+^ and CD8^+^ T-cell counts in subjects who recovered from severe COVID-19. Bars indicate median levels. *P* values are shown.

## DISCUSSION

A large body of evidence has accumulated on the features of SARS-CoV-2-specific adaptive immunity in patients who recovered from mild or severe clinical forms of COVID-19 [1-4,8-18,23]. To our knowledge, these studies mostly involved recently recovered patients (up to 3 months from onset of symptoms), of whom a large percentage (45-95%) consistently exhibited both T- and B-cell responses. The dynamics of such immune responses beyond this time point remains largely unexplored. Elucidating whether SARS-CoV-2-targeted T- and B-cell responses persist over time following natural infection, and if so for how long, is of paramount relevance from a clinical and public health perspective, mainly because it may allow us to predict susceptibility to reinfections upon re-exposure. The key issue to resolve is whether SARS-CoV-2 T- and B-cell memory responses are durable (over years), as may occur following SARS-CoV infection [19,20], or instead wane over a short period until undetectable, as seen in seasonal coronavirus infections [20].

Here, we assessed SARS-CoV-2 T- and B-cell immune responses in patients who had recovered from severe COVID-19 at medium term (from 2 to 6 months) after disease presentation. We used a whole blood flow cytometry ICS method for detection and enumeration of activated and functional (IFN-γ-producing) SARS-CoV-2-reactive CD4^+^ and CD8^+^ T cells, targeting the S1 region of the S protein and the M protein. Both proteins contain highly immunodominant HLA-I and HLA-II restricted epitopes eliciting readily detectable activated and/or functional T cells (CD4^+^ T cells more frequently), in most convalescent or short-term recovered COVID-19 patients [10-18]. Like other SARS-CoV-2 T-cell immunoassays [11,12,14,15], our ICS assay appears capable of quantifying coronavirus cross-reactive T cells, as two out of seven non-exposed individuals exhibited detectable responses (not shown).

Likewise, RBD-specific IgG levels strongly correlate with NtAb titers as measured using live or S-pseudotyped SARS-CoV-2, so they can be used as a proxy for inferring the neutralizing activity of sera [6,7,23].

Several major findings arose from our study. First, SARS-CoV-2-S1/M-reactive-IFN-γ CD4^+^ and CD8^+^ T cells were detected in a limited number of recovered patients from severe COVID-19 (around 30% and 10% for CD4^+^ and CD8^+^ T cells, respectively). Furthermore, we were unable to document SARS-CoV-2 T-cell reactivity beyond day 130 after COVID-19 diagnosis. This latter observation must be interpreted with caution given the low number of patients (n=3) tested within that timeframe. In a previous study also recruiting severe COVID-19 patients [21], we reported that around 40% of patients were capable of mounting SARS-CoV-2-S1/M-reactive IFN-γ-producing CD8^+^ T cell responses shortly after infection (median, 27 days after symptoms onset); in that study, SARS-CoV-2-reactive CD4^+^ T cell responses were not assessed. Nevertheless, other studies including hospitalized COVID-19 patients [11,12] found detectable functional CD4^+^ T-cell responses to both S1 and M proteins in a large fraction of patients either during the acute phase of the disease or at short-term convalescence. Taken collectively, the above observations suggest that functional SARS-CoV-2-S1/M-reactive T-cell frequencies in peripheral blood may wane over time. If confirmed in further studies, this phenomenon may be associated with COVID-19 severity. In this sense it has been postulated that exuberant or aberrant activation of the innate immune system upon encounter with SARS-CoV-2, resulting in suboptimal expansion of functional T cells, may be a hallmark in patients developing the severest forms of COVID-19 [1,25]. In support of this assumption, around 60% of individuals who had experienced mild forms of the disease were shown to display S1 and M-reactive T-cell responses by 6 months after COVID-19 diagnosis [26]. Likewise, Dan et al. [27] reported that SARS-CoV-2- (both S and M)-specific CD4^+^ and CD8^+^ T cells declined over time, with a half-life of 3-5 months in a cohort (n=41) skewed toward mild forms of COVID-19, although they remained detectable in many subjects at more than 6 months after COVID-19 diagnosis.

Second, in the current series, 60% of individuals displayed detectable SARS-CoV-2-RBD-specific IgGs at 2-5 months following COVID-19 diagnosis. In this context, it has been consistently reported that RBD IgG seroconversion occurs almost universally in moderate to severe COVID-19 patients within 3-4 weeks after onset of symptoms [6,7,22]. The above data thus suggest that the likelihood of detecting such antibody specificity in sera diminishes over time. Moreover, by analyzing paired serum specimens collected within the first month after symptoms onset and by 3-5 months after COVID-19 diagnosis in recovered individuals, we showed that the detection rate decreased slightly (from 94% to 77%) over time, as was the case of SARS-CoV-2-RBD-specific serum IgG levels.

Third, we found no correlation between SARS-CoV-2-reactive IFN-γ CD4^+^ T cells and RBD-specific antibody levels, suggesting that SARS-CoV-2 T- and B-cell responses may follow divergent kinetics, and perhaps that CD4^+^ T-cell help is not strictly required to sustain SARS-CoV-2-RBD-specific IgG responses over the medium term, at least in the population group analyzed herein. Other studies [10,28] found a strong correlation between NtAb antibody titers and the numbers of virus-specific T cells targeting S or N proteins in short-term convalescent individuals.

Fourth, frequency and logistic regression analyses identified presence of one or more comorbidities, but not age, sex or COVID-19 severity (ICU vs. non-ICU), as a factor presumably hampering the persistence of peripheral blood SARS-CoV-2-S1/M-reactive T cells in recovered COVID-19 patients. As for RBD IgGs, both sex (male) and COVID-19 severity appeared to increase the probability of detectable responses at medium term after disease presentation, although neither of these had an impact on RBD IgG levels.

Fifth, since uncontrolled inflammation driven by innate immune response shortly following viral infection may negatively affect the strength and durability of arising T-cell responses [23], we investigated whether serum peak levels of inflammatory biomarkers (IL-6, Dimer-D and ferritin) measured early after COVID-19 presentation (within 15 days) differed across recovered COVID-19 patients with or without detectable SARS-CoV-2-reactive T cells or SARS-COV-2-RBG IgGs, finding no variation. Moreover, little or no correlation was observed between the magnitude of systemic inflammation and SARS-CoV-2 immune parameters.

The current study has several limitations that must be acknowledged; predominantly, the reduced cohort size. Second, T-cell immunity analyses were conducted at a single time point. We cannot rule out that detectability of the two SARS-CoV-2-reactive T cells by the immunoassay used herein may fluctuate over time. Moreover, no whole blood specimens drawn at the time of patient hospitalization were available for T-cell immunity assessment. Third, T cells and antibodies targeting other antigen specificities, which may afford protection against SARS-CoV-2, were not evaluated. Fourth, T-cell functionalities other than IFN-γ production upon antigenic stimulation were not explored. Fifth, we cannot be certain whether our whole blood T-cell immunoassay might be less sensitive than others using fresh or cryopreserved isolated PBMCs. Sixth, taking serum peak levels of IL-6, Dimer-D and ferritin during the first 15 days after symptoms onset is an admittedly arbitrary time point for analyses which may not have captured the true net state of systemic inflammation generated shortly after SARS-CoV-2 infection.

In summary, we have shown that a relatively low number of subjects who developed severe forms of COVID-19 had detectable SARS-CoV-2-S1/M IFNγ CD4^+^ and CD8^+^ T cells at medium term after clinical diagnosis (up to 6 months), particularly those with concurrent comorbidities. Our data also indicated that serum levels of RBD-specific IgGs decline over time, becoming undetectable in some patients. Elucidating whether individuals lacking these specific adaptive immune responses are susceptible to reinfection is beyond the scope of the current study and needs to be addressed in future research.

## Data Availability

Provided data upon reasonable request.

## ACKNOWLEDGMENTS

We thank all personnel working at Clinic University Hospital for their unwavering commitment in the fight against COVID-19.

## FINANCIAL SUPPORT

This work received no public or private funds.

## CONFLICTS OF INTEREST

The authors declare no conflicts of interest.

## AUTHOR CONTRIBUTIONS

BO, EA, IT, PA, MJR and RG-R: Methodology and data validation. BO, EA, IT: Formal analysis. JR-D, JB, DN: Conceptualization, supervision. MLB, JR and JS-C: Clinical Staff in charge of patients. DN: writing the original draft. All authors reviewed and approved the original draft.

## FIGURE LEGENDS

**Supplementary Figure 1**. Enumeration of SARS-CoV-2-S1/M-reactive CD69^+^-expressing IFN-γ-producing CD4^+^ and CD8^+^ T cells by flow cytometry for intracellular staining in COVID-19 patients. (a) Lymphocyte gating; (b) CD3^+^ gating; CD8^+^ gating; (d) CD3^+^/CD69^+^/IFN-γCD8^+^/Isotype control; (e-f) CD3^+^/CD69^+^/IFN-γCD8^+^ positive individual; (g-h) CD3^+^/CD69^+^/IFN-γCD8^+^ negative individual. Dot-plot figures were built with FlowJo software (BD Biosciences).

## Notes

### Competing Interest Statement

The authors have declared no competing interest.

### Funding Statement

No external funding was received.

### Author Declarations

The current study was approved by the Research Ethics Committee of Hospital Clinico Universitario INCLIVA (March 2020)

