## Supplementary material for "Adaptive immune responses to SARS-CoV-2 in recovered severe COVID-19 patients": Table 1

| **TABLE 1. Detectable SARS-CoV-2-reactive T cells and RBD-specific IgGs in COVID-19 patients according to demographics and clinical factors** | | | | | | |
| --- | --- | --- | --- | --- | --- | --- |
| Parameter | **SARS-CoV-2 CD4^+^ or CD8^+^ T cell response** | | | **SARS-CoV-2 RBD-specific IgGs** | | |
|  | Yes | No | *P* value | Yes | No | *P* value |
| Sex: Male/female | 13/8 | 25/12 | 0.66 | 18/3 | 5/9 | 0.04 |
| Age: ≤62.5/>62.5 years^a^ | 13/8 | 16/21 | 0.07 | 9/12 | 7/7 | 0.67 |
| ICU admission: Yes/No | 8/13 | 13/24 | 0.82 | 13/8 | 3/11 | 0.019 |
| Comorbidities: Yes/No | 9/12 | 26/11 | 0.04 | 15/6 | 9/5 | 0.65 |
| ^a^Median age of patients.  RBD, receptor binding domain of S protein. | | | | | | |
