## Supplementary Table 1 for "Adaptive immune responses to SARS-CoV-2 in recovered severe COVID-19 patients"

| **Supplementary TABLE 1. Correlation between serum levels of inflammatory biomarkers and SARS-CoV-2 CD4^+^ or CD8^+^ T cells and SARS-CoV-RBD-specific IgGs** | | |
| --- | --- | --- |
| **Parameters** | **Spearman Rho value** | **P value** |
| IL-6/ SARS-CoV-2 CD4^+^ T cells | 0.07 | 0.68 |
| IL-6/ SARS-CoV-2 CD8^+^ T cells | 0.11 | 0.51 |
| IL-6/ SARS-CoV-2 RBD-IgGs | -0.01 | 0.96 |
| D-D/ SARS-CoV-2 CD4^+^ T cells | -0.10 | 0.46 |
| D-D/ SARS-CoV-2 CD8^+^ T cells | 0.20 | 0.12 |
| D-D/ SARS-CoV-2 RBD-IgGs | 0.4 | 0.01 |
| Ferritin/ SARS-CoV-2 CD4^+^ T cells | -0.04 | 0.76 |
| Ferritin/ SARS-CoV-2 CD8^+^ T cells | 0.16 | 0.24 |
| Ferritin/ SARS-CoV-2 RBD-IgGs | 0.34 | 0.06 |
| D-D, Dimer-D; IL-6, interleukin-6; RBD, receptor binding domain. | | |
